# COVID-19 in Pakistan: A national analysis of five pandemic waves

**DOI:** 10.1101/2023.01.23.23284902

**Authors:** Taimoor Ahmad, Mujahid Abdullah, Abdul Mueed, Faisal Sultan, Ayesha Khan, Adnan Ahmad Khan

**Affiliations:** Akhter Hameed Khan Foundation, Islamabad, Pakistan; Ministry of National Health Services, Regulation and Coordination, Islamabad, Pakistan; Shaukat Khanum Memorial Cancer Hospital & Research Centre, Lahore, Pakistan; Research and Development Solutions, Islamabad, Pakistan

## Abstract

**Objectives:** The COVID-19 pandemic showed distinct waves where cases ebbed and flowed. While each country had slight, nuanced differences, lessons from each wave with country-specific details provides important lessons for prevention, understanding medical outcomes and the role of vaccines. This paper compares key characteristics from the five different COVID-19 waves in Pakistan.

**Methods:** We used specific criteria to define COVID-19 waves, and key variables such as COVID-19 tests, cases, and deaths with their rates of change to the peak and then to the trough were used to draw descriptive comparisons. Additionally, a linear regression model estimated daily new COVID-19 deaths in Pakistan.

**Results:** Pakistan saw five distinct waves, each of which displayed the typical topology of a complete infectious disease epidemic. The time from wave-start to peak became progressively shorter, and from wave-peak to trough, progressively longer. Each wave appears to also be getting shorter, except for wave 4, which lasted longer than wave 3. A one percent increase in vaccinations increased daily new COVID-19 deaths by 0.10% (95% CI: 0.01, 0.20) in wave 4 and decreased deaths by 0.38% (95% CI: -0.67, -0.08) in wave 5.

**Conclusion:** Each wave displayed distinct characteristics that must be interpreted in the context of the level of response and the variant driving the epidemic. Key indicators suggest that COVID-19 preventive measures kept pace with the disease. Waves 1 and 2 were mainly about prevention and learning how to clinically manage patients. Vaccination started late during Wave 3 and its impact became apparent on hospitalizations and deaths in Wave 5. The impact of highly virulent strains Alpha/B1.1.7 and Delta/B.1.617.2 variants during Wave 3 and milder but more infectious Omicron/BA.5.2.1.7 are apparent.

## INTRODUCTION

In Pakistan, the first case of COVID-19, a novel and little-understood disease, was detected on February 26, 2020. Being a developing country with limited resources, crumbling health infrastructure and low health expenditure (1), Pakistan has no past experience with pandemics and a high burden of communicable diseases (2). As of February 23, 2022, the country had fully vaccinated 43% of its total population (3) and the Omicron variant of COVID-19 was the dominant strain (4). These factors make Pakistan a high-risk country, with a large pool of infection-susceptible people.

The emergence of COVID-19 has arguably been the biggest social and economic disruption in Pakistan in recent history. The pandemic has largely manifested itself in five distinct waves each of which have a rise, plateau and trough in cases, followed by a period of dormancy, after which the incidence of COVID-19 infections begins to rise again. Thus, each individual wave follows a four stage pattern followed by endemicity that has been seen for many infectious disease epidemics (12). Beyond anecdotal observation, there is evidence that this is happening with COVID-19 as well (11). What sets COVID-19 apart is that after completion of an individual wave, a new one would come along shortly, rather than much long, for example, annual, recurrences for influenza etc. This pattern has been seen all across the globe (5–10) with the timing of COVID-19 waves in different countries broadly coinciding (11).

In this context, current literature on COVID-19 largely focuses on high-income countries during the initial waves (2,13–17), or aggregated at a regional level (5,6). Given the different capacities of countries to manage the pandemic (17), there is a need to explore the characteristics of the subsequent pandemic waves in a developing country context, preferably with granularity of a country-level analysis (2,13–17).

To this end, this paper explores how each of the five waves of the COVID-19 pandemic in Pakistan differed from each other in terms of the total duration of each wave, the time from the start of each wave to its peak, and from its peak to the wave’s trough; the length of time in between each wave; as well as key descriptive features like the total number of COVID-19 tests, COVID-19 cases, hospitalizations, deaths from COVID-19, and COVID-19 vaccinations in each wave. Additionally, this study statistically models the key factors that lead to deaths from COVID-19.

## METHODS

### Criteria for COVID-19 waves

We begin by retrospectively defining various time periods between 2020 and 2022 as distinct waves, based on existing literature (18). There are a total of 628 observations (daily set of indicators) across these five waves. Based on our criteria, the starting point of each COVID-19 wave is defined as the day with the lowest number of daily new COVID-19 cases preceding a consistent rise in these cases, before the peak of the respective COVID-19 wave. The end of each wave is defined as the day with lowest number of daily new COVID-19 cases following a 7-day decline; this number also needed to be lower than the cases on any of the 7 days that followed it (Table 1).

**Table 1.** Augmented Dicky-Fuller tests for unit roots.

| Variable | Test Statistics | P-Value | H<sub>0</sub>: Random Walk |
| --- | --- | --- | --- |
| Daily New COVID-19 Deaths | -6.96 | 0.000 | Stationary |
| Daily New COVID-19 Cases | -3.23 | 0.018 | Stationary |
| Daily New COVID-19 Tests | -3.94 | 0.0017 | Stationary |
| Oxford Containment and Health Index | -3.30 | 0.015 | Stationary |
| Ventilator-admitted Ratio | -5.69 | 0.000 | Stationary |
| Oxygen-Admitted Ratio | -3.00 | 0.034 | Stationary |
| Fully Vaccinated People | -2.77 | 0.003 | Stationary |

### Data and variables

In order to estimate the pattern for COVID-19 throughout the five waves in Pakistan, we have used time series of various daily indicators from April 3, 2020 to February 23, 2022, which categorized into following themes:

i. Wave timespan
ii. COVID-19 tests
iii. COVID-19 cases
iv. Test-to-case ratio
v. COVID-19 positivity
vi. Hospitalization, treatment, and containment
vii. COVID-19 deaths
viii. COVID-19 vaccination
ix. Policy environment

Several variables in the list above were transformed into ratios for the purpose of describing all five COVID-19 waves in Pakistan (Supplementary Table S1).

The data for all but two of the above variables, second doses of COVID-19 vaccines administered and the Oxford Containment and Health Index for COVID-19, was compiled from daily national situation reports (Sitreps). These Sitreps are prepared by the National Emergency Operations Centre (NEOC) in Islamabad, Pakistan. Data in these Sitreps have served as the basis for all major COVID-19 policy decisions in Pakistan.

The data for second doses of COVID-19 vaccines administered was sourced directly from the National Command & Operation Centre (NCOC), Islamabad, Pakistan, which is the government forum that brings together the ministries of Health and Planning along with the military to determine pandemic policy and to coordinate the response. Data for the Oxford Containment and Health Index is taken from a publicly available dataset from the University of Oxford’s Blavatnik School of Governance (19). This dataset is compiled by using qualitative information about the non-pharmaceutical interventions (NPIs) in a country and quantifying them into an index. A detailed methodology of the index calculation can be found in a working paper by the Blavatnik school (20).

### Model Specification

For our model for COVID-19 deaths, we used a linear ordinary least square (OLS) regression carried out using STATA 17. The data as well as the model was divided into five distinct periods, representing the five waves of COVID-19 in Pakistan, as of February 2022. Regression analysis was performed separately on each wave and results were compared with each other to make logical inferences.

Our dependent variable is the Daily New COVID-19 Deaths in Pakistan in logarithmic form (**LnY**_**t**_); ϵ_t_ represents the error term capturing the effect of omitted variables. The specification of our linear OLS regression model is as following

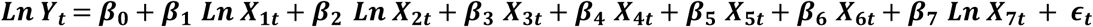

We have chosen seven independent variables:

i. Log of daily new COVID-19 cases with 21-day delay (**LnX**_**1t**_);
ii. Log of daily new COVID-19 tests with 28-day delay (**LnX**_**2t**_);
iii. The Oxford Containment and Health Index for COVID-19 with 14-day delay (**X**_**3t**_);
iv. Time (**X**_**4t**_);
v. The number of people on ventilators as a proportion of the total admitted (**X**_**5t**_);
vi. The number of people on oxygen as a portion of the total admitted (**X**_**6t**_);
vii. Log of second doses of COVID-19 vaccines administered with 14-day delay (**LnX**_**7t**_)

Daily new COVID-19 cases were regressed with a 21-day lag, since among those who die from COVID-19 infection, death occurs between a median of 14 days (21) and 25 days (average of three weeks) after presenting symptoms (22,23). This is pertinent in the case of Pakistan, as most of the COVID-19 testing in the country has been symptomatic, i.e., done when someone develops symptoms of COVID-19 and hence either voluntarily gets tested or is prescribed by a medical professional to do so.

Given the delay for daily new cases, daily new COVID-19 tests were regressed with a delay of 28 days. This delay allows for the time it takes for someone to test positive for COVID-19 and for their symptoms to worsen (for example, by escalating to hospitalization, which takes nearly a week (24) before resulting in death. For vaccination, a 14-day lag was taken, as immunity from vaccines is generally understood to develop two weeks or longer after receiving a shot (25–27).

The Oxford Containment and Health Index is calculated out of 100 which means strict restrictions and a lower value indicate a smaller number of restrictions imposed on the general population. This variable was regressed with a 14-day lag, as we assume that any new government restrictions will take approximately that long to have any effect. Additionally, the time variable is meant to capture any unmeasured or seasonal effects on COVID-19 deaths in Pakistan, such as an overall rate of increase/ decrease of daily deaths in each wave. We assume the error term is not correlated with any of the independent variable.

Diagnostic tests were performed on the estimated model to ensure the precision of the estimated coefficient: for heteroskedasticity, the Breusch-Pagan test was applied, whereas for serial correlation, the Durbin-Watson test was used. Variance inflation factor (VIF) was calculated for multicollinearity. Dickey-Fuller tests were run for each independent variable in our regression model. All of the variables were found to be stationary, fulfilling an important pre-requisite for our analysis (Table 1).

## RESULTS

### Summary statistics

Pakistan experienced five distinct waves from 3^rd^ April 2020 till 23^rd^ February 2022 (Fig 1). The first wave lasted the longest (150 days), while the fifth one was the shortest (83 days). The fourth wave was remarkable for its relatively rapid upslope and a long tail, while the fifth wave showed the reverse pattern. The duration of each wave of COVID-19 in Pakistan was shorter than the preceding one apart from wave 4. After wave 1, each wave took less time to reach its peak and took longer to reach its trough apart from wave 5.

**Figure 1.**
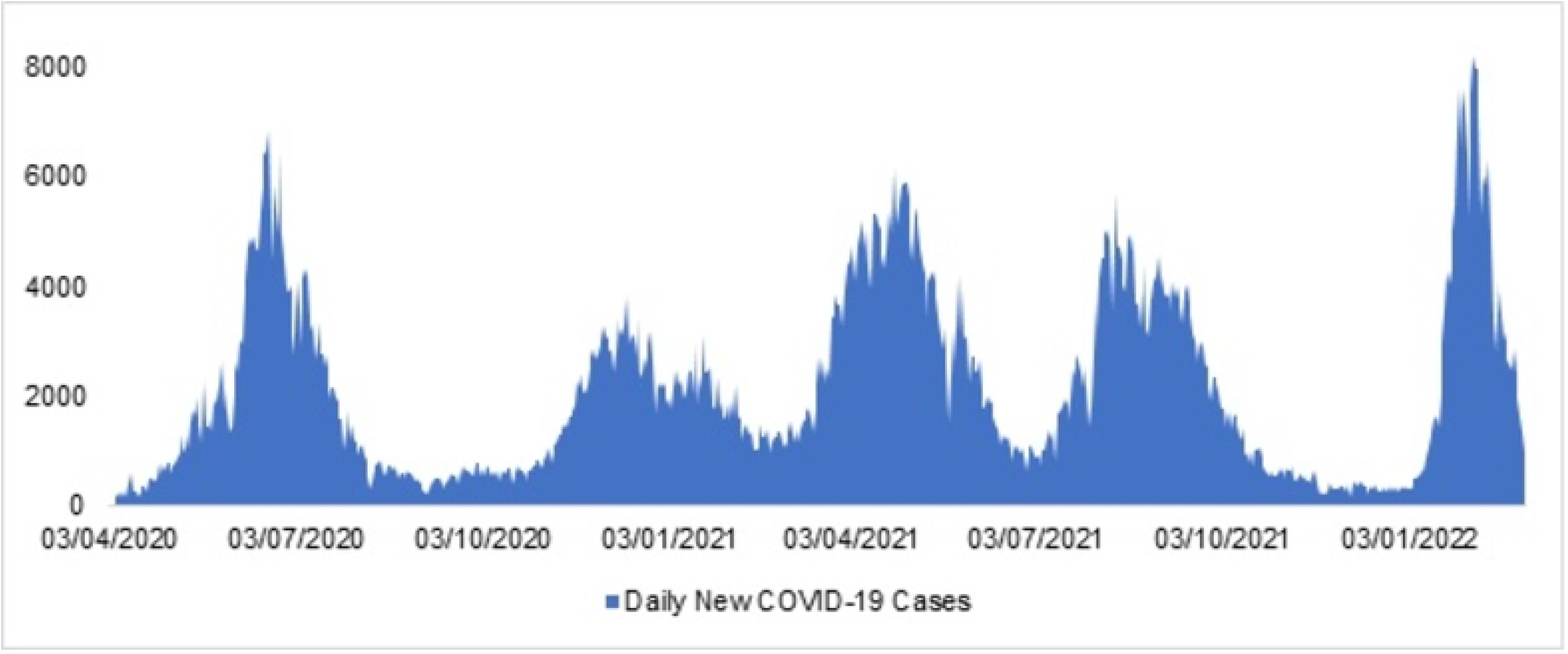
COVID-19 waves and daily new cases.

The capacity to conduct tests expanded over time from an average of 17,142 tests daily during the first wave to 49,650 during fifth wave. The increase in the daily tests peaked during the fourth wave. The highest average daily number of cases (3147) were observed during the wave 3. The rate of increase of COVID-19 cases was highest during wave 4, but the rate of decline in cases after the peak of a wave was fastest during wave 5. Test to case ratio kept on increasing from 15 during the first wave to 57 during the fifth one. While total positivity varied across waves, the rate in daily change of positivity remained relatively unchanged apart from wave 2 and 3, where it is lower as compared to other waves (Table 2).

**Table 2.**
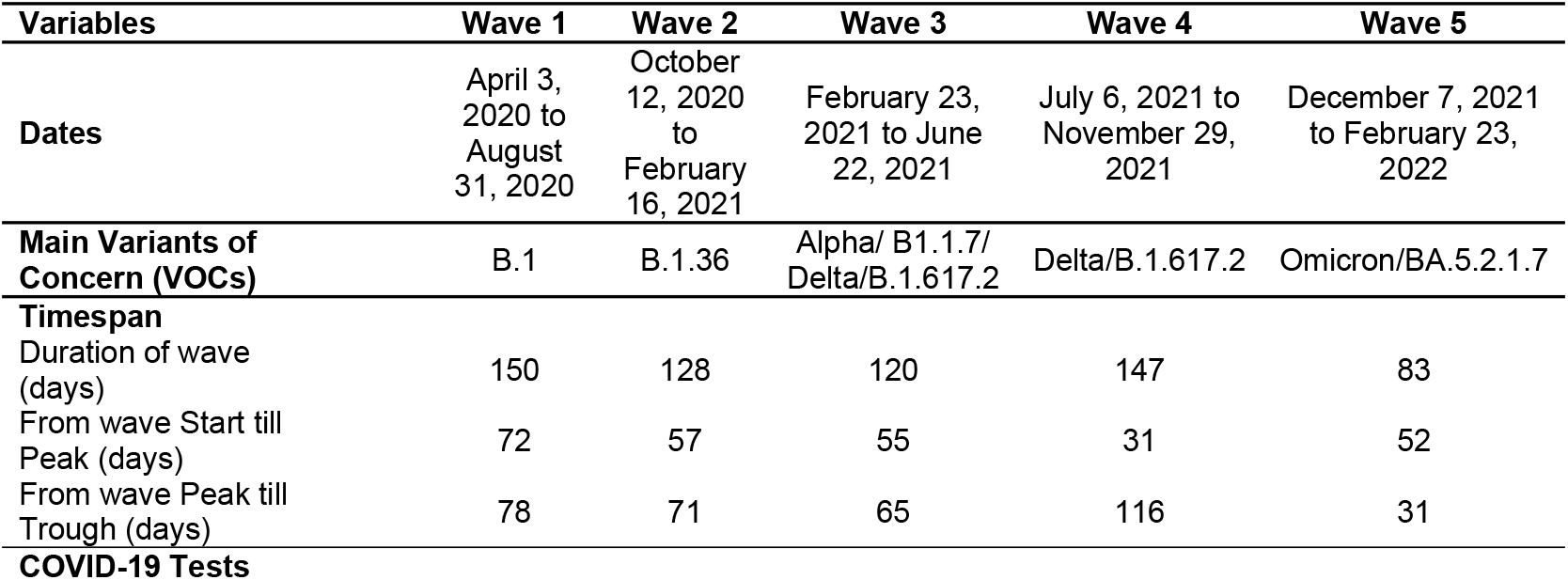
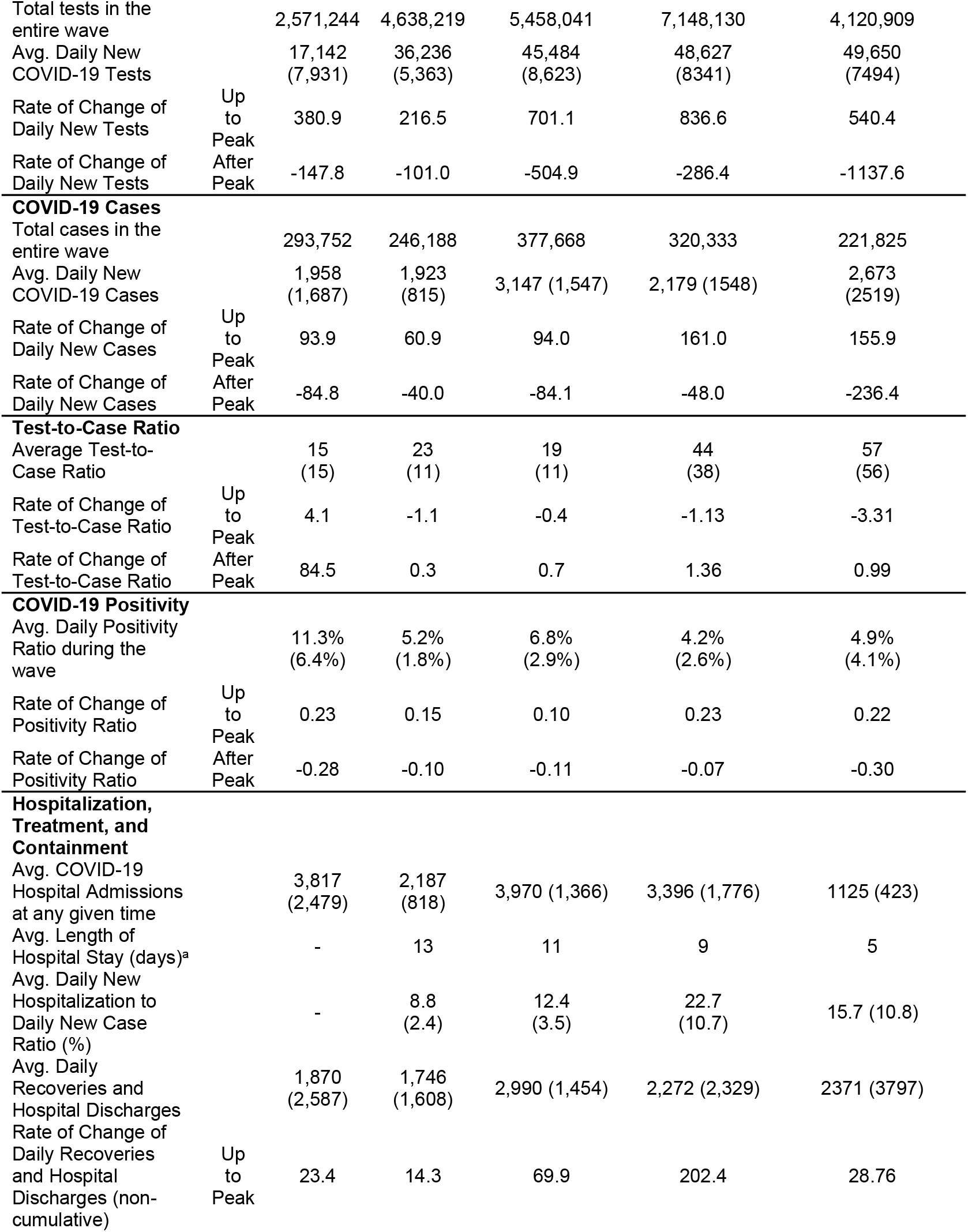
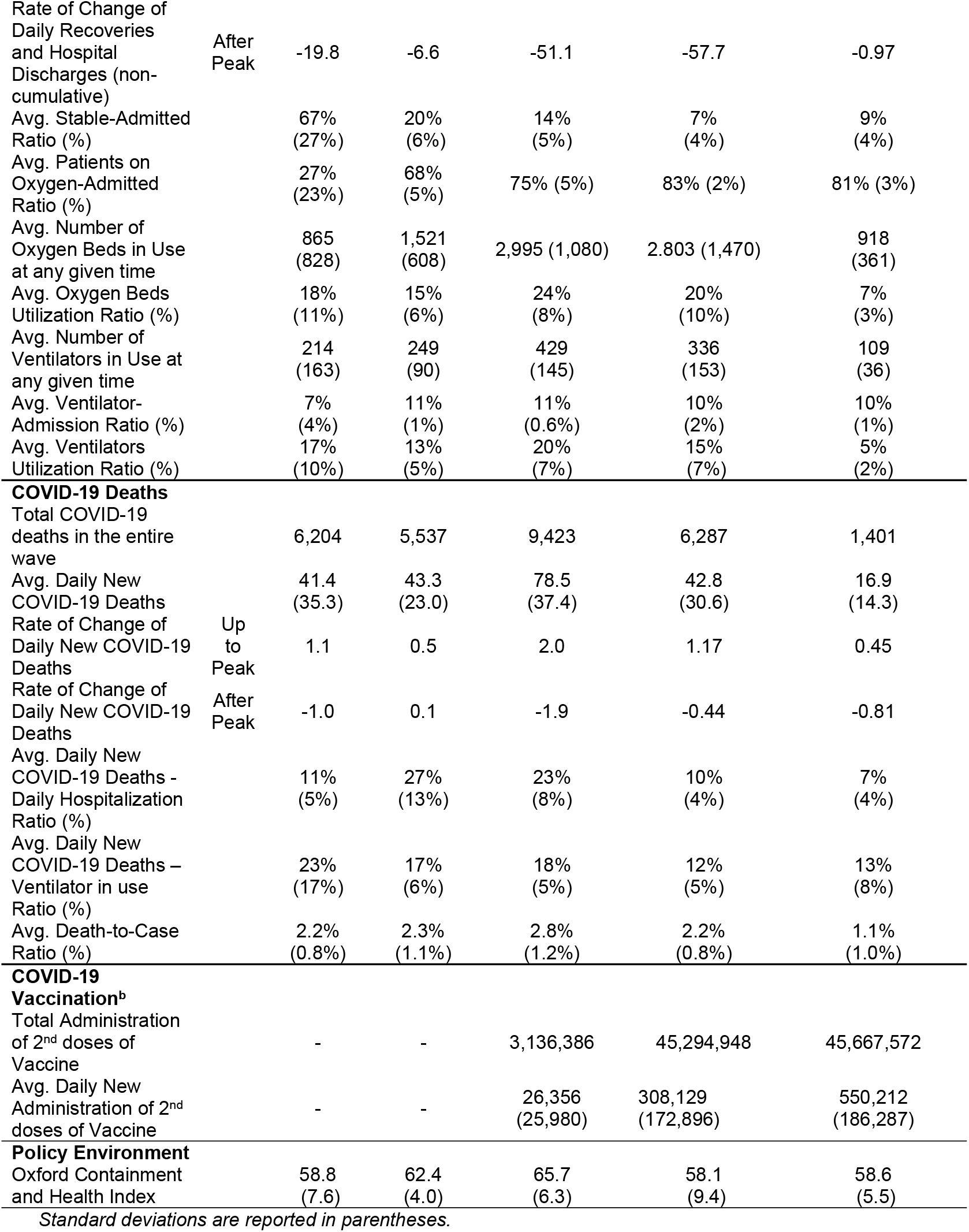
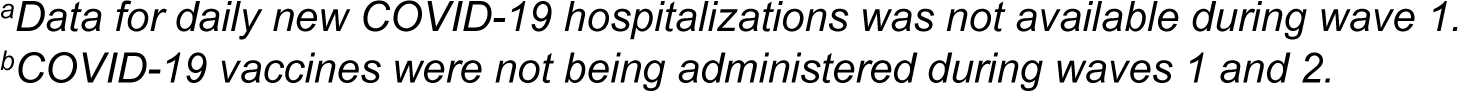
Descriptive characteristics of each COVID-19 wave.

Hospitalizations were the highest for the first and the third wave and the least for the fifth, whereas duration of hospitalization fell linearly from an initial 13 days during the second wave to 5 days during the fifth. Hospitalizations became more specific over time in that, nearly two third of admitted patients during the first wave were stable, compared to 9% during the fifth one. The average stable-admitted ratio decreased continuously from wave 1 to wave 4 but increased slightly in wave 5. The rate at which people recovered from COVID-19 and/or were discharged from hospital was highest in wave 4 and slowest in wave 2.

The average Oxygen Beds-Admitted Ratio continuously increasing in each wave, reaching its maximum value in wave 4. During the first wave 27% of all admissions required oxygen, 7% needed a ventilator, compared to 81% and 10% respectively during the fifth wave. The average oxygen bed utilization followed a declining trend except for the third wave (24%) and was lowest in the fifth wave (7%). The average Ventilators Utilization Ratio shows that all available ventilators were not fully utilized in any of the five waves. The highest ratio was in wave 3 (20%) and the lowest ratio was in wave 5 (5%). These two ratios suggest that most critical patients were put on oxygen for recovery and a small proportion of these people were transferred to ventilators.

Deaths from COVID-19 were the highest during the third wave at 9,423, which also saw the highest daily number of deaths (78.5) and the highest rate of increase in daily deaths. Average daily deaths to hospitalization peaked during the second wave, while deaths to ventilator use was the highest during the first. Average death to case ratio was the highest for the third wave but was in the 2.2-2.8% range, with the exception of the fifth wave when it was 1.1%.

Pakistan’s vaccination drive started towards the end of wave 2, but full vaccination (i.e., people receiving both their doses) did not occur until the beginning of wave 3. Consequently, total and daily new second dose of vaccine administered was highest in wave 5. Government restrictions, measured by the Oxford Health and Containment Index, appear to have been comparable in each wave.

## Regression results

The results indicate the daily new COVID-19 cases were a statistically significant determinant for daily new deaths in all five waves at 95% CI; a one-percentage increase in COVID-19 cases caused a 0.46-0.69 % increase in deaths across the five waves (Table 3).

**Table 3.**
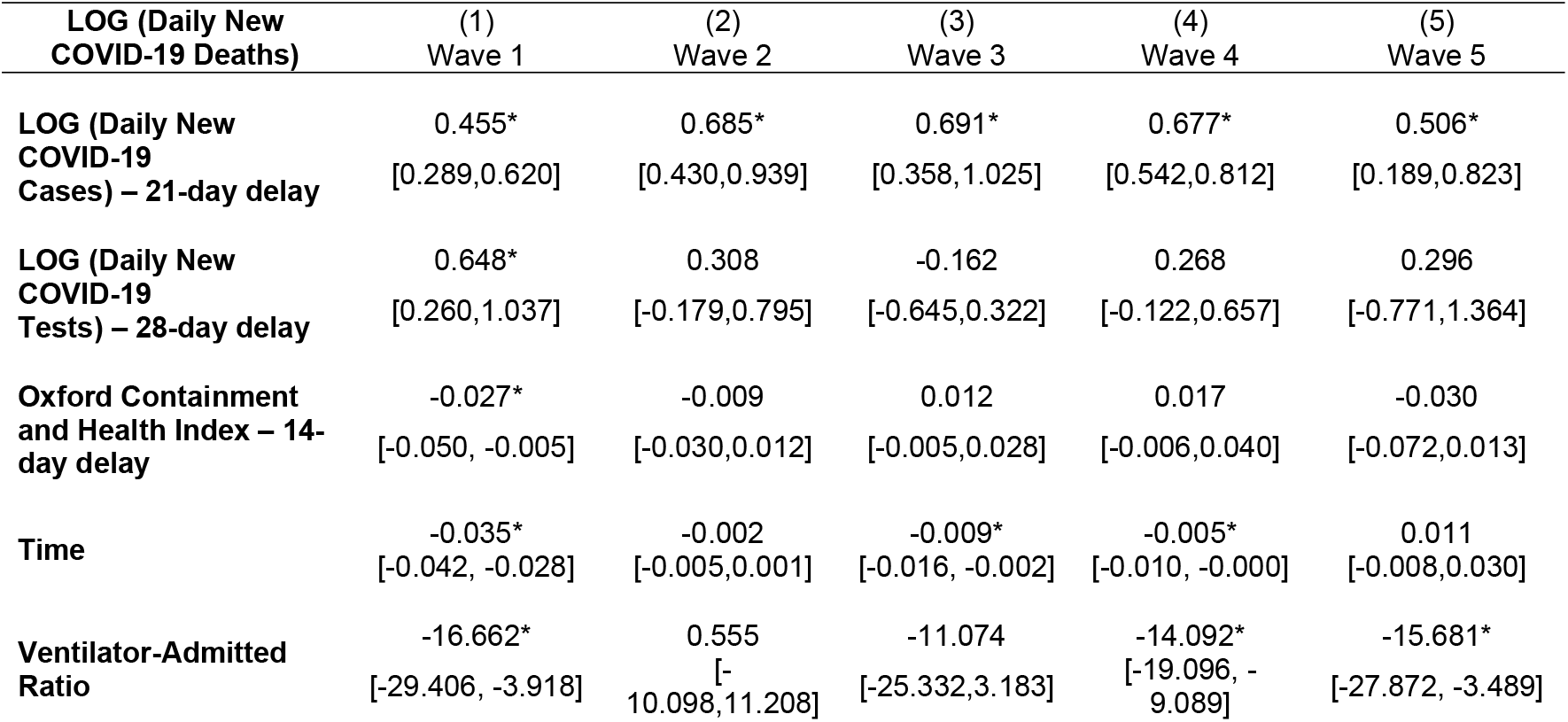
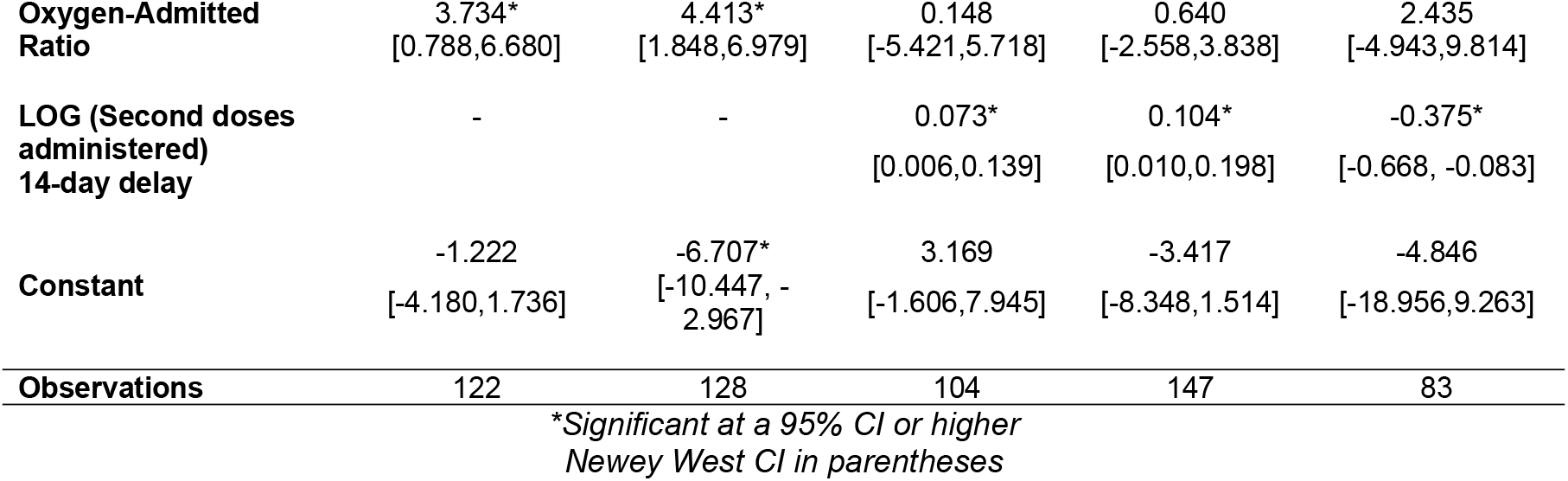
Ordinary least squares (OLS) regression results for COVID-19 deaths.

The daily new COVID-19 tests and Oxford Health and Containment Index, which records the presence of government NPIs and restrictions, were both found to be a statistically significant determinants of COVID-19 deaths in wave 1. Increasing daily new tests by 1% will reduce the daily deaths by 0.65% (95% CI: 0.26%, 1.04%). While increase in Oxford Health and Containment Index by 1 point will result in 0.03% reduction in daily new deaths (95% CI: -0.05%, -0.005%).

The time trend variable is statistically significant in wave 1, wave 3 and wave 4. The coefficients indicate that, on average, daily COVID-19 deaths decreased at a rate of 0.04% per day (95% CI: -0.04%, -0.03%) as time passed during wave 1. However, daily new deaths reduced at a rate of 0.01% per day during wave 3 (95% CI: -0.02%, 0.002%) and wave 4 (95% CI: -0.01%, 0.00%).

The ventilator to admitted ratio was statistically significant in wave 1, wave 4 and wave 5. The coefficient remains negative throughout these three waves and significant at 95% CI. The coefficients indicates that if ratio of patients on ventilator out of the admitted increases than daily new deaths decrease by 14-17%.

Oxygen to admitted ratio is only significant in wave 1 and 2 at 95% CI, where the coefficient is positive. It means that if the ratio of oxygenated patient out of the total admitted increase than daily new deaths due to COVID-19 will increase by approximately 4%.

Lastly, the number of fully vaccinated people came out to be statistically significant in each of the last three waves. During wave 3, COVID-19 deaths increase by 0.07% (95% CI: 0.006,0.14) as percentage of fully vaccinated people increase by one addition percentage. This rate increased to 0.10% (95% CI: 0.006,0.14) during the wave 4. However, in the fifth wave, daily new deaths due to COVID-19 decrease by 0.38% (95% CI: -0.67, -0.08) as fully vaccinated people increases.

## DISCUSSION

Pakistan experienced 5 distinct waves from 3^rd^ April 2020 to 23^rd^ February 2022. Our analysis reflects both the evolution of Pakistan’s response, as well as the differential impact of different variants of the virus shaped the contours and features of each wave. Pakistan experienced its initial wave earlier than other South Asian countries including India, Bangladesh, Sri Lanka and Nepal, while peaks for the subsequent waves coincided with those in other countries (31).

The upslope, as seen by the rate of change for testing and cases, was always steeper than during the downward slope of a wave. This pattern follows what is known about infectious epidemics in that cases rise quickly, plateau and then fall, slowly to an endemic state where a low ebb of infections persist in the community in definitely (12). In fact, each wave behaved as a typical epidemic caused by a distinct variant of the virus. Wave 1 was dominated by B.1 variant, wave 2 by B.1.36 variant, the wave 3 by Alpha/B1.1.7 and Delta/B.1.617.2 variants, wave 4 had majority cases of the Delta/B.1.617.2 variant (28– 30) while wave 5 was driven by Omicron/BA.5.2.1.7 (4).

A key challenge faced by Pakistan at the beginning of the pandemic was that there was little prior experience with any pandemic outbreak of such level. Although disease surveillance systems exist, they had not been scaled to manage case surveillance, hospital admissions, daily deaths, and eventually large-scale adult vaccination and event tracking. Pakistan has a federal system of governance where provinces provide health services while the federal ministry provides guidance and coordination. In addition, considerable curative care is in the private sector. To address the potential difficulties in mounting a unified national response to the disease in the face of this diversity, a National Action Plan for COVID-19 was formulated in March 2020 that placed the responsibility for the national response in a National Coordination Committee (NCC) that was headed by the Prime Minister and attended by all federal ministers. The NCC set national policy which was implemented by the National Command and Operation Centre (NCOC) that was co-headed by the military and civilian leadership (32). The NCOC coordinated the management of the extensive lockdowns, other key NPIs such as school closures, limited opening hours for essential businesses (examples of which included grocery stores and pharmacies), closure of borders, cancellation of public events and social gatherings (33,34). This was supported based on an elaborate data gathering and analysis system that that guided daily decisions.

Wave 1 continued the longest and intervals became shorter between each successive wave. Each wave showed unique features, that were determined by the particular variant that drove that wave, along with the larger context that included the type of the variant driving the wave, the extent and type of preventive interventions and eventually the availability of the vaccine.

Pakistan’s response to COVID-19 evolved over time. For example, Wave 1 had the highest positivity rates and the longest duration, in part due to low initial rates of testing, including very little contact tracing in the early days (35). As testing increased and mobility restrictions tightened, duration of Waves 2 and 3 became shorter. However, by the end of the second wave, intervention fatigue had set in. Implementation was laxer, and these factors contributed to more cases and deaths of any wave during Wave 3. Indeed, the Oxford Health and Containment Index was significant only during Wave 1 in terms of preventing deaths.

In addition to preventive measures, the higher daily COVID-19 cases in waves 3, 4 and 5 may be attributed to highly transmissible Alpha (36,37), Delta (38,39) and Omicron (40) variants, and to easing of severe restrictions such as lockdowns and school closures (41). It is also possible that many cases were missed during Wave 1 due to limited testing. However, the stability of daily testing in waves 3 to 5 suggests a stable equilibrium between the testing system and how cases were being incident – the system was capturing most of the cases from previously recognized populations and locations. It is likely that undiagnosed cases and deaths were few, since as part of the national surveillance, teams kept abreast of burials in large and midsized towns and also periodically canvased opinion of general practitioners about upsurges in respiratory illnesses. On average, Pakistan had fewer cases per million population than neighboring countries of India, Bangladesh, and Iran, as well as several of the developed countries (31).

As with prevention, clinical management of cases evolved over time. Initially most cases were hospitalized as seen by the high case to hospitalization ratio – only 27% of admissions required oxygen 7% required ventilators during the Wave 1. In fact, there was a correlation between deaths and oxygenation (which was mostly at hospitals) during waves 1 and 2, a pattern that was seen globally. However, with each succeeding wave, use of oxygen increased while ventilators fluctuated within a narrow range, as was also seen in India (52,53). Thus, even as COVID-19 hospitalizations peaked during Wave 3, hospitalization to case ratio increased, and average duration of hospitalization and the use of hospitals for simple oxygenation fell, suggesting hospitals, ICU and ventilators, were increasingly reserved only for the sickest (42). Deaths correlated best with a 21-day delay model rather than a 28-day one, suggesting that most deaths happened early after infection. Higher hospitalizations during the third wave may also have been attributed to the Alpha followed by Delta variants (43–45). By contrast the lower hospitalizations, length of stay and mortality during the fifth wave may be attributed to the Omicron variant that was seen worldwide (46,47), and specifically in South Africa (48) and Brazil (49) during the Omicron waves. Vaccination started halfway during wave 3 and more than half of the eligible population was fully vaccinated (3) by the fifth wave, and may have contributed to lower hospitalizations in Wave 5. Unlike COVID-19 induced major challenges to the healthcare capacity in various countries (50,51), Pakistan was able to build healthcare resources capacity to keep pace with the pandemic. Ventilator and oxygen utilization never exceeded 20% and 24% respectively in wave 3.

Vaccination drive started in Pakistan by the end of February 2021. Despite a slow start, vaccination picked up pace from 26,356 daily vaccinations in Wave 3 to 308,129 in the fourth wave as it was rolled-out to younger population and vaccine supply increased in the country. Average daily deaths did not reduce significantly due to vaccinations during the third and fourth waves (56,57) but showed marked reduction in hospitalizations and deaths towards the end of fourth and during the entire fifth wave (58).

From our regression model, we found that daily new COVID-19 cases were statistically significant determinants of daily new deaths due to COVID-19. The association was also observed from the third wave as both cumulative cases and deaths were the highest, including the average daily deaths which were considerably higher than any other wave, as seen in other countries (54). Secondly, daily new deaths due to COVID-19 increased with patients on oxygenated beds while decreased with patients on ventilators in the initial waves, potentially due to high patients load in hospitals, critical patients were put on oxygen rather than ventilator. Fifth wave experienced the smallest number of daily COVID-19 deaths possibly because it was dominated by the Omicron variant (55).

## LIMITATIONS

There are a number of limitations associated with the data used in this paper. While the official data used for the analysis are disaggregated by sub-national level, demographic disaggregation, such as age or gender, are not available. This limits the analysis in terms of the implication of gender and age on COVID-19 deaths. The national data is compiled by aggregating the numbers for each subnational unit in Pakistan. However, such an analysis would be too extensive to depict and therefore our analysis does not account for sub-national differences. It is possible that distinctive cultures, behaviors and differences in the stringency in enforcement of interventions vary between for regions and may in theory, influence the number of COVID-19 cases and deaths.

Similarly, data for hospitalizations is also unaccompanied by any information on comorbidities, as this information was not available beyond treating hospitals, losing a level of richness of analysis that includes such comorbidities. Also, data for daily new hospital admissions started becoming available towards the very end of wave 1. Consequently, the average length of hospital stay could not be calculated for this wave.

The official vaccination data available to us at the time of this analysis is not desegregated by the different types of available vaccines, for example Sinopharm, CanSino, Sputnik V and others. Differential impact of each vaccine on COVID-19 deaths in Pakistan would be informative. All of the above limitations notwithstanding, we are confident that this study provides crucial insights into the prevailing trends of COVID-19 in Pakistan in manner that is constructive.

## CONCLUSION

COVID-19 management in Pakistan kept pace with the spread of the disease during five distinct waves. We describe how these waves differed in terms of cases, hospitalizations and deaths and analyze potential reasons for these differences. These lessons, as well as limitations of our approach, provide important lessons for managing future pandemics.

## Data Availability

The data was provided to the Akhter Hameed Khan Foundation (AHK-F) team for this study as part of its work with Pakistan’s Federal Ministry of National Health Services, Regulations & Coordination (MoNHSR&C) and the National Command & Operation Centre (NCOC) in Islamabad, which are leading Pakistan’s response to the COVID-19 pandemic. The AHK-F team has provided analytical support to the above entities, and such created knowledge that has directly informed pandemic policy-making in Pakistan. COVID-19 data is compiled and shared in daily National Situation Reports, or Sitreps, by the National Emergency Operation Centre (NEOC). Each day’s Sitrep is compiled as a PDF file. The data used for this study was manually compiled from these PDF files and then used in STATA. The parentage of this data is with the NCOC and the MoNHSR&C. The AHK-F team received this data with the express understanding that it would be kept confidential. However, the data can be obtained independently from the NEOC, through a data request procedure, which is subject to approval from the MoNHSR&C. The data request itself is to be addressed to: Dr. Shahzad Baig, National Coordinator, National Emergency & Operation Center, D Block, EPI Building, Chak Shahzad, Park Road, Islamabad. Phone: +92-51-8730879. The data on Oxford Health and Containment Index is taken from and publicly available at the following GitHub repository: https://github.com/OxCGRT/covid-policy-tracker/tree/master/data

## List of Legends

**S1 Table. Calculated ratio variables and their descriptions**

